# EPIDEMIOLOGY OF ANAPHYLAXIS IN COLOMBIA 2010-2015

**DOI:** 10.1101/2023.06.27.23291967

**Authors:** Pablo Andrés Miranda-Machado

## Abstract

**Background:** The lifetime prevalence of anaphylaxis from all triggers is estimated to range from 0.05 % to 2.6 %. Data on anaphylaxis in the Latin American region and Colombia are scant.

**Objective:** Estimate the prevalence, incidence and mortality for anaphylaxis in Colombia.

**Methods:** Records with diagnosis of anaphylaxis (ICD-10 code T78.0-T78.2) of Information System of Social Protection (SISPRO) between 2010 and 2015 were included. To determine the prevalence, incidence and mortality for anaphylaxis, population estimates from the National Statistics Department of Colombia (DANE) were used.

**Results:** The average estimated annual prevalence of anaphylaxis were 25.1 cases per million. The average estimated annual incidence of anaphylaxis were 3.2 cases per million. Most cases range between 45 and 49 years age. On average 8.4 cases of anaphylaxis deaths per year were estimated. The average estimated annual mortality of anaphylaxis were 0.16 cases per million.

**Conclusions:** Both under-diagnosis and under-report of anaphylaxis are common in the world. Population studies in Colombia, with diagnoses of based primarily on clinical history and clinical criteria for accurate and early identification of anaphylaxis recently established are required.

## INTRODUCTION

Anaphylaxis, a term first used by Richet and Portier in 1902, is defined as an immediate systemic or generalized hypersensitivity reaction, mediated or not by immunoglobulin E (IgE), with release of pro-inflammatory mediators by mast cells and basophils (1). It is a potentially lethal reaction that affects multiple organs or systems, and with a sudden and unexpected onset, which can progress from mild symptoms to a very serious and occasionally irreversible reaction (2). Beta-lactam antibiotics from the penicillin group, analgesics from the group of non-steroidal anti-inflammatory drugs (NSAIDs) and food are the main causes of anaphylaxis due to IgE-dependent immunological mechanisms. Intravenous iodinated contrast media, iron dextran and blood products are the main causes of anaphylaxis due to IgE-independent immunological mechanisms.

The lifetime prevalence of anaphylaxis caused by or associated with any trigger or immunological mechanism is at least 1%. The incidence is unknown and mortality is rare, predominantly in asthmatics, adolescents, and long-lived adults with multiple comorbidities. The epidemiology of anaphylaxis in many countries, including Colombia, is unknown. Some studies have reported food, medicines, and insect bites as the main causes of anaphylaxis in Latin America (3).

According to the Latin American Online Survey on Anaphylaxis (OLASA), conducted between 2008 and 2010 in 15 Latin American countries and Portugal, 58.4% of patients who developed anaphylaxis were women and 68.5% were older than 18 years (5). The real incidence of anaphylaxis is estimated in a range of 10 to 20 cases per 100,000 people per year (4–6). In Europe, an incidence of anaphylaxis is estimated to be between 1.5-7.9 cases per 100,000 people per year and in more recent studies, a global incidence of anaphylaxis is estimated to be between 50-112 episodes per 100,000 people each year (9). Severe anaphylaxis affects between 10-30 people per 100,000 per year and causes death in 0.65-2% of patients who develop anaphylaxis (8). In registries of voluntary declarations of deaths, government databases and other studies, mortality from anaphylaxis is estimated to be between 0.33-3 deaths per million people per year (5,9).

The objective of this study was to estimate the prevalence, incidence, and mortality from anaphylaxis in Colombia between 2010 and 2015, in order to contribute to the epidemiological estimates of this immune-allergic problem and to describe the general panorama of this potentially lethal and preventable pathology.

## METHODS

A descriptive observational study of a cross-sectional cohort of data from patients diagnosed with anaphylaxis in Colombia was carried out during the period between 2010 and 2015. The objectives of this study were to estimate the prevalence, incidence, and mortality from anaphylaxis in Colombia during the period of study. The records of patients with a diagnosis of anaphylaxis registered in the Data Warehouse of the Social Protection Information System (SISPRO) between 2010 and 2015, who met the inclusion and exclusion criteria, were included. The inclusion criteria were the data of the patients with a diagnosis of anaphylaxis (ICD-10 code T78.0-T78.2) in the SISPRO between 2010 and 2015. The data of the patients with age, sex and type of diagnosis not reported or not defined were excluded. Prevalence, incidence, and mortality from anaphylaxis were estimated using the population projections from the 2005 Census of the National Administrative Department of Statistics (DANE) in Colombia. To estimate the prevalence, data from patients with all types of diagnoses (diagnostic impression, new confirmed, and repeated confirmed) were taken into account. For the estimation of the incidences, the data of the patients with a new type of confirmed diagnosis were taken into account. For the estimation of mortality, the data of patients with exit status (death) were taken into account.

The information in the SISPRO Data Warehouse contains all the Individual Health Service Provision Records (IHSPR) of all the authorized Healthcare Provider Institutions (HPI) in Colombia. This information was obtained with the authorization of SISPRO prior induction, training and access to the server cubos.sispro.gov.co. DANE information is public and was consulted on the institution’s website (www.dane.gov.co).

### Statistic Analysis

The variables will be summarized in means +/- standard deviations for the quantitative variables and in percentages for the qualitative variables. For normally distributed variables, means +/- standard deviations will be reported. The univariate analysis by age and sex to search for an association with the diagnosis of anaphylaxis was performed using the Chi Square statistic. A p value < 0.05 was considered significant.

## RESULTS

5,829 cases with anaphylaxis were identified between 2010 and 2015 (59.1% women). 745 new confirmed cases were confirmed in the same period (49.6% women). On average, 149 new cases of anaphylaxis per year were estimated. The mean estimated annual prevalence of anaphylaxis was 25.1 cases per million (2010 = 11.5; 2011 = 11.5; 2012 = 11.4; 2013 = 5.4; 2014 = 14.1 and 2015 = 11.9) (**Table 1-3**).

**Table 1.** Cases treated with diagnosis of anaphylaxis 2010-2015. Source: SISPRO.

| **Ages** | **2010** | **2011** | **2012** | **2013** | **2014** | **2015** | **Total** |
| --- | --- | --- | --- | --- | --- | --- | --- |
| De 0 a 4 years | 45 | 38 | 32 | 194 | 55 | 47 | 405 |
| De 05 a 09 years | 29 | 24 | 20 | 145 | 40 | 34 | 286 |
| De 10 a 14 years | 40 | 38 | 27 | 120 | 33 | 26 | 278 |
| De 15 a 19 years | 39 | 39 | 36 | 156 | 57 | 34 | 355 |
| De 20 a 24 years | 41 | 37 | 39 | 220 | 49 | 38 | 422 |
| De 25 a 29 years | 36 | 47 | 41 | 259 | 54 | 49 | 484 |
| De 30 a 34 years | 44 | 40 | 49 | 301 | 51 | 50 | 530 |
| De 35 a 39 years | 32 | 31 | 41 | 232 | 56 | 38 | 426 |
| De 40 a 44 years | 39 | 28 | 42 | 239 | 37 | 41 | 419 |
| De 45 a 49 years | 50 | 46 | 42 | 253 | 51 | 47 | 480 |
| De 50 a 54 years | 34 | 46 | 39 | 249 | 38 | 45 | 448 |
| De 55 a 59 years | 28 | 33 | 40 | 192 | 44 | 28 | 360 |
| De 60 a 64 years | 19 | 30 | 33 | 155 | 43 | 31 | 303 |
| De 65 a 69 years | 16 | 15 | 26 | 119 | 27 | 24 | 227 |
| De 70 a 74 years | 10 | 14 | 11 | 95 | 26 | 9 | 165 |
| De 75 a 79 years | 13 | 14 | 8 | 78 | 10 | 12 | 135 |
| De 80 years or older | 11 | 13 | 16 | 80 | 6 | 23 | 149 |
| **Total** | **523** | **529** | **532** | **3,083** | **673** | **575** | **5,829** |

**Table 2.** Male cases treated with anaphylaxis diagnosis 2010-2015. Source: SISPRO.

| **Ages** | **2010** | **2011** | **2012** | **2013** | **2014** | **2015** | **Total** |
| --- | --- | --- | --- | --- | --- | --- | --- |
| De 0 a 4 years | 19 | 25 | 18 | 96 | 31 | 27 | 211 |
| De 05 a 09 years | 16 | 12 | 8 | 73 | 20 | 15 | 138 |
| De 10 a 14 years | 23 | 26 | 19 | 57 | 13 | 18 | 151 |
| De 15 a 19 years | 18 | 21 | 20 | 57 | 29 | 19 | 164 |
| De 20 a 24 years | 19 | 14 | 19 | 53 | 21 | 17 | 143 |
| De 25 a 29 years | 17 | 17 | 20 | 60 | 19 | 23 | 156 |
| De 30 a 34 years | 22 | 13 | 25 | 82 | 24 | 16 | 180 |
| De 35 a 39 years | 12 | 17 | 15 | 70 | 22 | 17 | 151 |
| De 40 a 44 years | 23 | 16 | 19 | 63 | 21 | 19 | 159 |
| De 45 a 49 years | 19 | 16 | 25 | 82 | 22 | 17 | 176 |
| De 50 a 54 years | 18 | 20 | 19 | 78 | 17 | 22 | 172 |
| De 55 a 59 years | 12 | 13 | 18 | 68 | 18 | 12 | 140 |
| De 60 a 64 years | 8 | 12 | 14 | 67 | 21 | 13 | 131 |
| De 65 a 69 years | 9 | 7 | 10 | 50 | 9 | 10 | 95 |
| De 70 a 74 years | 4 | 4 | 5 | 44 | 14 | 2 | 73 |
| De 75 a 79 years | 6 | 5 | 4 | 34 | 7 | 6 | 62 |
| De 80 years or older | 6 | 6 | 5 | 35 |  | 13 | 65 |
| **Total** | **249** | **244** | **256** | **1,066** | **306** | **266** | **2,344** |

**Table 3.** Female cases treated with anaphylaxis diagnosis 2010-2015. Source: SISPRO.

| Ages | 2010 | 2011 | 2012 | 2013 | 2014 | 2015 | Total |
| --- | --- | --- | --- | --- | --- | --- | --- |
| De 0 a 4 years | 23 | 12 | 13 | 97 | 24 | 20 | 188 |
| De 05 a 09 years | 11 | 9 | 11 | 65 | 19 | 19 | 134 |
| De 10 a 14 years | 16 | 11 | 6 | 56 | 18 | 6 | 112 |
| De 15 a 19 years | 20 | 18 | 14 | 97 | 28 | 14 | 187 |
| De 20 a 24 years | 22 | 23 | 20 | 167 | 28 | 21 | 279 |
| De 25 a 29 years | 19 | 30 | 21 | 199 | 35 | 26 | 328 |
| De 30 a 34 years | 22 | 27 | 24 | 219 | 27 | 33 | 349 |
| De 35 a 39 years | 20 | 14 | 26 | 162 | 34 | 21 | 275 |
| De 40 a 44 years | 16 | 12 | 23 | 176 | 16 | 22 | 260 |
| De 45 a 49 years | 31 | 30 | 17 | 171 | 29 | 29 | 303 |
| De 50 a 54 years | 16 | 26 | 20 | 171 | 21 | 23 | 276 |
| De 55 a 59 years | 16 | 20 | 22 | 124 | 26 | 16 | 220 |
| De 60 a 64 years | 11 | 18 | 19 | 88 | 22 | 18 | 172 |
| De 65 a 69 years | 7 | 8 | 16 | 69 | 18 | 14 | 132 |
| De 70 a 74 years | 6 | 10 | 6 | 51 | 12 | 7 | 92 |
| De 75 a 79 years | 7 | 9 | 4 | 44 | 3 | 6 | 73 |
| De 80 years or older | 5 | 7 | 11 | 45 | 6 | 10 | 84 |
| **Total** | **268** | **280** | **270** | **2,000** | **364** | **304** | **3,445** |

The estimated mean annual incidence of anaphylaxis was 3.2 cases per million (2010 = 2.9; 2011 = 2.9; 2012 = 2.5; 2013 = 2.3; 2014 = 3.0 and 2015 = 2.5). Most of the cases ranged between 45 and 49 years (**Table 4-6**).

**Table 4.** New cases attended with diagnosis of anaphylaxis 2010-2015. Source: SISPRO.

| Ages | 2010 | 2011 | 2012 | 2013 | 2014 | 2015 | Total |
| --- | --- | --- | --- | --- | --- | --- | --- |
| De 0 a 4 years | 13 | 10 | 9 | 11 | 11 | 12 | 66 |
| De 05 a 09 years | 11 | 9 | 2 | 3 | 9 | 9 | 43 |
| De 10 a 14 years | 10 | 7 | 8 | 5 | 6 | 4 | 39 |
| De 15 a 19 years | 12 | 14 | 7 | 8 | 11 | 8 | 58 |
| De 20 a 24 years | 9 | 10 | 6 | 6 | 14 | 7 | 51 |
| De 25 a 29 years | 8 | 7 | 9 | 7 | 12 | 8 | 51 |
| De 30 a 34 years | 13 | 4 | 11 | 5 | 8 | 7 | 47 |
| De 35 a 39 years | 8 | 9 | 12 | 9 | 12 | 7 | 57 |
| De 40 a 44 years | 11 | 6 | 9 | 11 | 10 | 12 | 58 |
| De 45 a 49 years | 9 | 15 | 11 | 12 | 8 | 9 | 62 |
| De 50 a 54 years | 9 | 12 | 10 | 9 | 5 | 9 | 53 |
| De 55 a 59 years | 7 | 10 | 9 | 8 | 9 | 6 | 48 |
| De 60 a 64 years | 1 | 8 | 8 | 4 | 12 | 10 | 43 |
| De 65 a 69 years | 5 | 3 | 3 | 4 | 5 | 7 | 27 |
| De 70 a 74 years | 4 | 5 | 2 | 7 | 5 | 3 | 26 |
| De 75 a 79 years | 3 | 1 | 3 | 1 | 4 |  | 12 |
| De 80 years or older | 1 | 5 | 2 |  | 2 | 4 | 14 |
| **Total** | **132** | **135** | **118** | **109** | **141** | **122** | **745** |

**Table 5.**
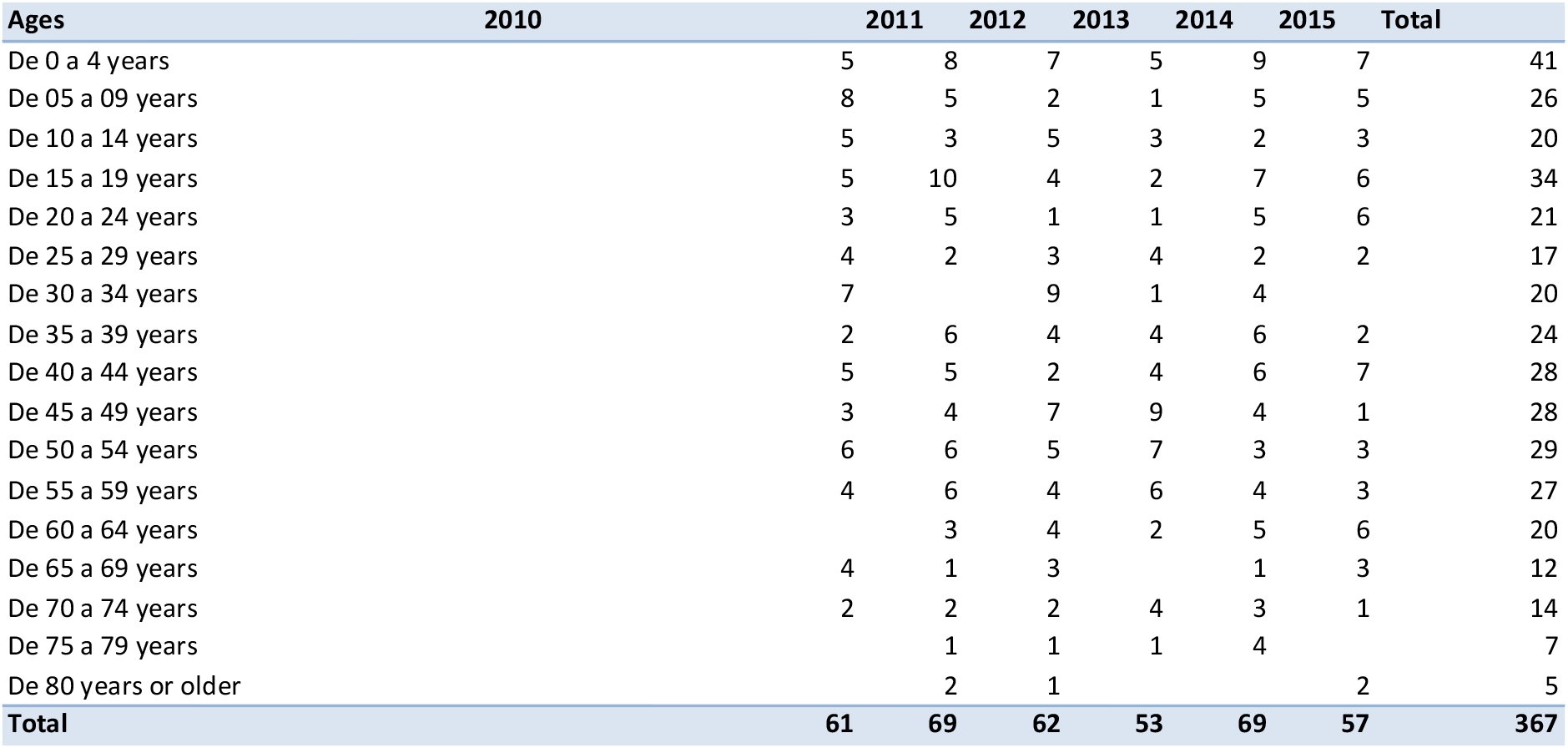
New male cases treated with anaphylaxis diagnosis 2010-2015. Source: SISPRO.

**Table 6.** New female cases treated with anaphylaxis diagnosis 2010-2015. Source: SISPRO.

| Ages | 2010 | 2011 | 2012 | 2013 | 2014 | 2015 | Total |
| --- | --- | --- | --- | --- | --- | --- | --- |
| De 0 a 4 years | 7 | 2 | 2 | 6 | 2 | 5 | 24 |
| De 05 a 09 years | 2 | 3 |  | 1 | 4 | 4 | 14 |
| De 10 a 14 years | 5 | 4 | 2 |  | 4 | 1 | 16 |
| De 15 a 19 years | 6 | 4 | 2 | 5 | 4 | 2 | 23 |
| De 20 a 24 years | 6 | 5 | 5 | 5 | 9 | 1 | 30 |
| De 25 a 29 years | 4 | 5 | 6 | 3 | 10 | 6 | 34 |
| De 30 a 34 years | 6 | 4 | 2 | 4 | 4 | 7 | 27 |
| De 35 a 39 years | 6 | 3 | 8 | 5 | 6 | 5 | 33 |
| De 40 a 44 years | 6 | 1 | 7 | 7 | 4 | 5 | 30 |
| De 45 a 49 years | 6 | 11 | 4 | 3 | 4 | 8 | 34 |
| De 50 a 54 years | 3 | 6 | 5 | 2 | 2 | 6 | 24 |
| De 55 a 59 years | 3 | 4 | 5 | 2 | 5 | 3 | 21 |
| De 60 a 64 years | 1 | 5 | 4 | 2 | 7 | 4 | 23 |
| De 65 a 69 years | 1 | 2 |  | 4 | 4 | 4 | 15 |
| De 70 a 74 years | 2 | 3 |  | 3 | 2 | 2 | 12 |
| De 75 a 79 years | 3 |  | 2 |  |  |  | 5 |
| De 80 years or older | 1 | 3 | 1 |  | 2 | 2 | 9 |
| Total | 68 | 65 | 54 | 52 | 72 | 65 | 370 |

On average, 8.4 cases of deaths due to anaphylaxis per year were estimated. Mean estimated annual mortality from anaphylaxis was 0.16 cases per million (2010 = 0.2; 2011 = 0.1; 2012 = 0.1; 2013 = 0.2; 2014 = 0.1 and 2015 = 0.1) (**Table 7-9**).

**Table 7.**
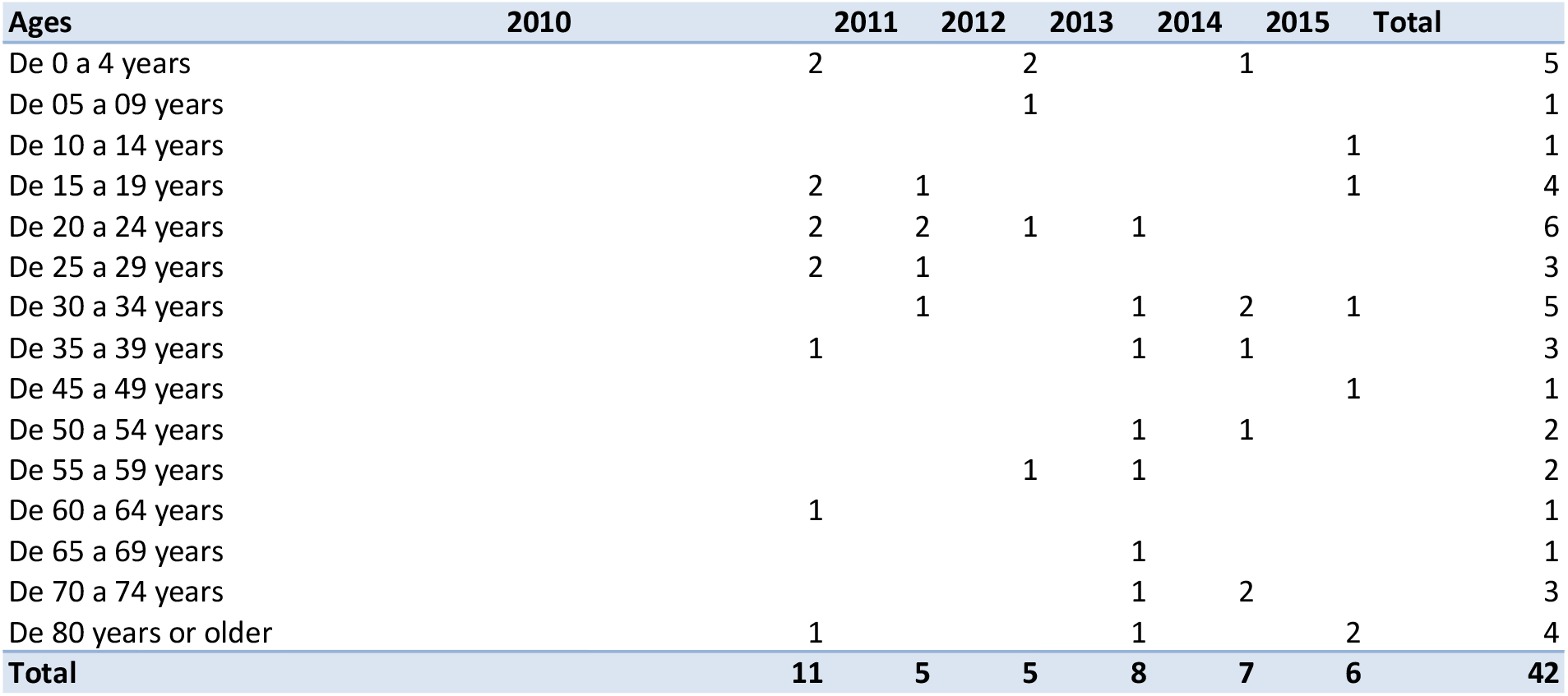
Deaths related to diagnosis of anaphylaxis 2010-2015. Source: SISPRO.

**Table 8.**
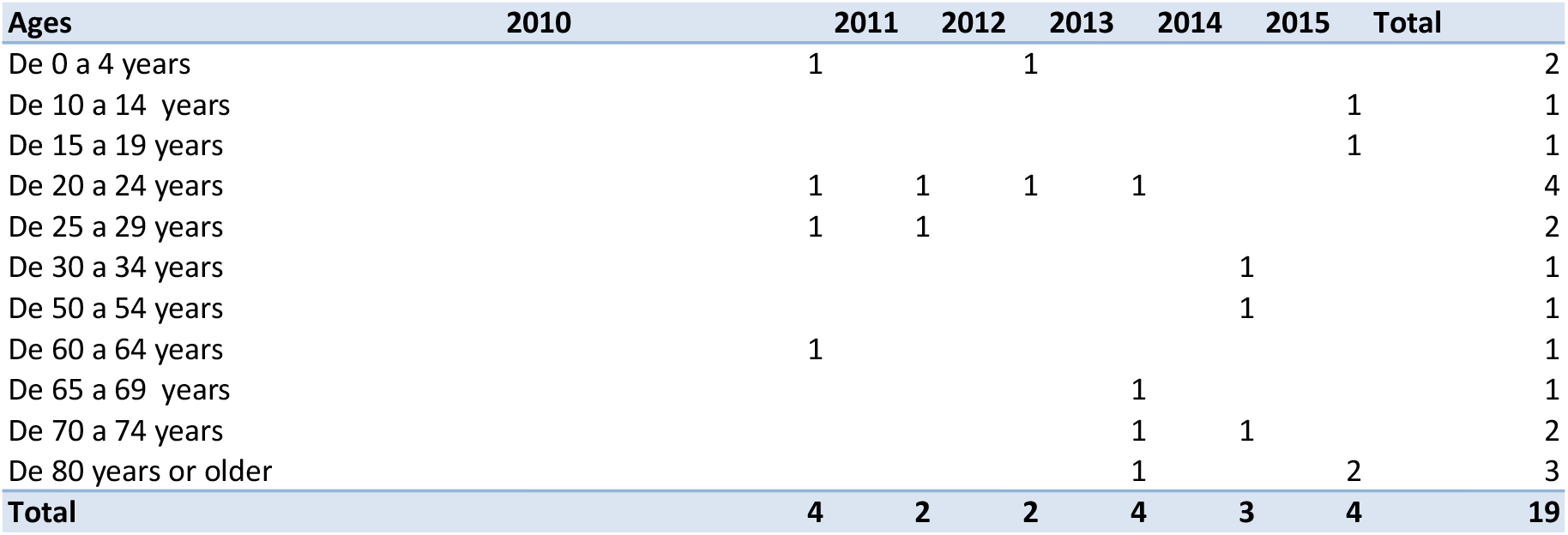
Male deaths related to anaphylaxis diagnosis 2010-2015. Source: SISPRO.

**Table 9.**
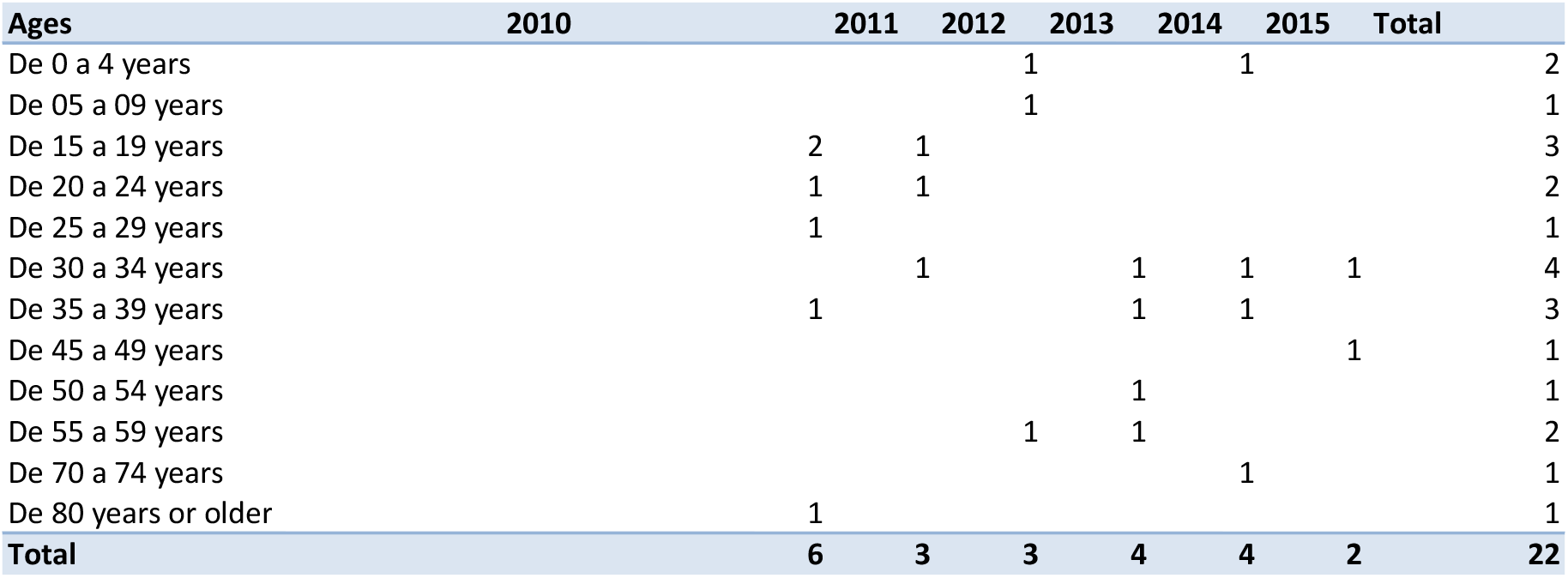
Female deaths related to anaphylaxis diagnosis 2010-2015. Source: SISPRO.

The Colombian population was taken from the population projections of the 2005 Census of DANE (**Table 10-12**).

**Table 10.** Population projections of Colombia 2010-2015. Source: DANE.

| POPULATION |  |  |  |  |  |  |
| --- | --- | --- | --- | --- | --- | --- |
| Total | 2010 | 2011 | 2012 | 2013 | 2014 | 2015 |
| De 0 a 4 years | 4279721 | 4284237 | 4291149 | 4299725 | 4310123 | 4321637 |
| De 05 a 09 years | 4308667 | 4286387 | 4272011 | 4264594 | 4260992 | 4258678 |
| De 10 a 14 years | 4424204 | 4393148 | 4359830 | 4327970 | 4301474 | 4282708 |
| De 15 a 19 years | 2252800 | 2257296 | 2253817 | 2244449 | 2231913 | 2218821 |
| De 20 a 24 years | 4048318 | 4110318 | 4169560 | 4222511 | 4264257 | 4292291 |
| De 25 a 29 years | 3615319 | 3678614 | 3746936 | 3818440 | 3889676 | 3957939 |
| De 30 a 34 years | 3266313 | 3323648 | 3375368 | 3425261 | 3478846 | 3539724 |
| De 35 a 39 years | 2927032 | 2956138 | 3007266 | 3073074 | 3141854 | 3205979 |
| De 40 a 44 years | 2933196 | 2923885 | 2903042 | 2880514 | 2869549 | 2879410 |
| De 45 a 49 years | 2737901 | 2790367 | 2832007 | 2862194 | 2879512 | 2883795 |
| De 50 a 54 years | 2288314 | 2374840 | 2460771 | 2542977 | 2617116 | 2680490 |
| De 55 a 59 years | 1815503 | 1890811 | 1969508 | 2051081 | 2134484 | 2218791 |
| De 60 a 64 years | 1412231 | 1473107 | 1533411 | 1594650 | 1659236 | 1728396 |
| De 65 a 69 years | 1048429 | 1089641 | 1139440 | 1194296 | 1250825 | 1307382 |
| De 70 a 74 years | 830113 | 837870 | 848094 | 865218 | 891781 | 926841 |
| De 75 a 79 years | 588245 | 615521 | 641029 | 660523 | 674766 | 684618 |
| De 80 years or older | 593662 | 612255 | 630983 | 650106 | 669643 | 689614 |
| Adults | 29835859 | 30419638 | 31006092 | 31590796 | 32168770 | 32737024 |
| Total | 45509584 | 46044601 | 46581823 | 47121089 | 47661787 | 48203405 |

**Table 11.** Colombian Female Population Projections 2010-2015. Source: DANE.

| Female | 2010 | 2011 | 2012 | 2013 | 2014 | 2015 |
| --- | --- | --- | --- | --- | --- | --- |
| De 0 a 4 years | 2188787 | 2191282 | 2195042 | 2199694 | 2205140 | 2211071 |
| De 05 a 09 years | 2108294 | 2096617 | 2088961 | 2084913 | 2082905 | 2081546 |
| De 10 a 14 years | 2165482 | 2150935 | 2134514 | 2118395 | 2104802 | 2095089 |
| De 15 a 19 years | 2139616 | 2146518 | 2147601 | 2143506 | 2135740 | 2126291 |
| De 20 a 24 years | 1988818 | 2012617 | 2037584 | 2061652 | 2081630 | 2095681 |
| De 25 a 29 years | 1835414 | 1858523 | 1881166 | 1904075 | 1927690 | 1952203 |
| De 30 a 34 years | 1674449 | 1703188 | 1729487 | 1754172 | 1778575 | 1803602 |
| De 35 a 39 years | 1514236 | 1527133 | 1551280 | 1583059 | 1616660 | 1648373 |
| De 40 a 44 years | 1528816 | 1523614 | 1511796 | 1498588 | 1491010 | 1494077 |
| De 45 a 49 years | 1427983 | 1456005 | 1478857 | 1495820 | 1505806 | 1508394 |
| De 50 a 54 years | 1199794 | 1245488 | 1290257 | 1332829 | 1371402 | 1404887 |
| De 55 a 59 years | 949986 | 991535 | 1035219 | 1080402 | 1126190 | 1171877 |
| De 60 a 64 years | 740840 | 774122 | 807167 | 840857 | 876569 | 915085 |
| De 65 a 69 years | 555546 | 577898 | 605242 | 635567 | 666944 | 698532 |
| De 70 a 74 years | 451857 | 456464 | 461941 | 471078 | 485493 | 504881 |
| De 75 a 79 years | 327970 | 344937 | 360717 | 372754 | 381431 | 387074 |
| De 80 years or older | 342889 | 354753 | 366840 | 379352 | 392287 | 405568 |
| Adults | 15381930 | 15675049 | 15969909 | 16264125 | 16555248 | 16841895 |
| Total | 23042924 | 23313302 | 23584736 | 23857050 | 24130117 | 24403726 |

**Table 12.** Colombian Male Population Projections 2010-2015. Source: DANE.

| Male | 2010 | 2011 | 2012 | 2013 | 2014 | 2015 |
| --- | --- | --- | --- | --- | --- | --- |
| De 0 a 4 years | 2090934 | 2092955 | 2096107 | 2100031 | 2104983 | 2110566 |
| De 05 a 09 years | 2200373 | 2189770 | 2183050 | 2179681 | 2178087 | 2177132 |
| De 10 a 14 years | 2258722 | 2242213 | 2225316 | 2209575 | 2196672 | 2187619 |
| De 15 a 19 years | 2252800 | 2257296 | 2253817 | 2244449 | 2231913 | 2218821 |
| De 20 a 24 years | 2059500 | 2097701 | 2131976 | 2160859 | 2182627 | 2196610 |
| De 25 a 29 years | 1779905 | 1820091 | 1865770 | 1914365 | 1961986 | 2005736 |
| De 30 a 34 years | 1591864 | 1620460 | 1645881 | 1671089 | 1700271 | 1736122 |
| De 35 a 39 years | 1412796 | 1429005 | 1455986 | 1490015 | 1525194 | 1557606 |
| De 40 a 44 years | 1404380 | 1400271 | 1391246 | 1381926 | 1378539 | 1385333 |
| De 45 a 49 years | 1309918 | 1334362 | 1353150 | 1366374 | 1373706 | 1375401 |
| De 50 a 54 years | 1088520 | 1129352 | 1170514 | 1210148 | 1245714 | 1275603 |
| De 55 a 59 years | 865517 | 899276 | 934289 | 970679 | 1008294 | 1046914 |
| De 60 a 64 years | 671391 | 698985 | 726244 | 753793 | 782667 | 813311 |
| De 65 a 69 years | 492883 | 511743 | 534198 | 558729 | 583881 | 608850 |
| De 70 a 74 years | 378256 | 381406 | 386153 | 394140 | 406288 | 421960 |
| De 75 a 79 years | 260275 | 270584 | 280312 | 287769 | 293335 | 297544 |
| De 80 years or older | 250773 | 257502 | 264143 | 270754 | 277356 | 284046 |
| Adults | 14453929 | 14744589 | 15036183 | 15326671 | 15613522 | 15895129 |
| Total | 22466660 | 22731299 | 22997087 | 23264039 | 23531670 | 23799679 |

## DISCUSSION

The prevalence of anaphylaxis is underestimated or underdiagnosed due to different factors such as discrepancies in the concept or definition of the problem and the difficulty to identify it as it is a process of acute clinical expression in the context of clinical and forensic medical scenarios. In our study, the estimated prevalence for Colombia was 0.002% per year. This prevalence is lower than that reported globally. The lifetime prevalence of anaphylaxis taking any trigger into account is estimated to be at least 1% (4).

It was estimated that the prevalence of anaphylaxis in Colombia was higher in women (59.1% of cases between 2010-2015). According to the Latin American Online Survey on Anaphylaxis (OLASA), conducted between 2008 and 2010 in 15 Latin American countries and Portugal, 58.4% of patients who developed anaphylaxis were women (5).

In this study, we estimated an incidence of anaphylaxis of 3.2 cases per million per year. This is much lower than the actual incidence of anaphylaxis, which is estimated to be in the range of 10 to 20 cases per 100,000 people per year (4–6). In Europe, an incidence of anaphylaxis is estimated between 1.5-7.9 cases per 100,000 people year and in more recent studies, a global incidence of anaphylaxis is estimated between 50-112 episodes per 100,000 people each year (9).

The estimated mortality from anaphylaxis for Colombia was 0.16 cases per million per year. Severe anaphylaxis affects between 10-30 people per 100,000 per year and causes death in 0.65-2% of patients who develop anaphylaxis (8). In registries of voluntary declarations of deaths, government databases and other studies, mortality from anaphylaxis is estimated to be between 0.33-3 deaths per million people per year (5,9).

This study has several limitations. To identify the cases of anaphylaxis, the ICD-10 codes (T78.0-T78.2) were used and there was no access to the medical records, paraclinical results and treatments received by the patients, which constitutes an important limitation for the estimates of the frequency estimators of anaphylaxis and to describe the clinical characteristics of the cases. We only had access to the two main diagnoses registered due to limitations of SISPRO and the population data used are from the 2005 Colombian Census of DANE, which could explain the underestimation of the data compared to the global data.

## CONCLUSIONS

Both underdiagnosis and underreporting of anaphylaxis are common in the world. Studies in the Colombian population are required, with diagnoses based mainly on the clinical history and clinical criteria for the accurate and early identification of recently established anaphylaxis.

## Ethical Responsibilities

### Protection of people and animals

The authors declare that no experiments were carried out on humans or animals for this research.

### Data confidentiality

The authors declare that no patient data appears in this article.

### Right to privacy and informed consent

The authors declare that no patient data appears in this article.

